# Associations between occupations and the occurrence of sarcomas: results of the French population-based case-control study ETIOSARC

**DOI:** 10.64898/2026.07.20.26358458

**Authors:** Céline Gramond, Léa Guillemin, Gaëlle Coureau, Thomas Systchenko, Karima Hammas, Antonia Gasch Illescas, Patricia Delafosse, Jean-Yves Blay, Françoise Ducimetière, Nicolas Penel, Maud Toulmonde, François Le Loarer, Gonzague de Pinieux, Isabelle Baldi, Alain Monnereau, Aude Lacourt, Simone Mathoulin-Pelissier, Brice Amadeo

**Affiliations:** Univ. Bordeaux, INSERM, BPH, U1219, EPICENE team, F-33000 Bordeaux, France; Registre des Cancers de Gironde, CHU de Bordeaux, F-33000 Bordeaux, France; Registre des Cancers Poitou-Charentes, CIC-INSERM1402, CHU de Poitiers, F-86000 Poitiers, France; Registre des Cancers du Haut-Rhin, Groupe Hospitalier de la Région de Mulhouse et Sud Alsace, F-68000 Mulhouse, France; Registre Général des Cancers de Lille et de sa Région, GCS Alliance Cancer, F-59000 Lille, France; Registre des Cancers de l’Isère, CHU de Grenoble, F-38000 Grenoble, France; Département d’Oncologie Médicale, Centre Léon Bérard, Université Claude Bernard Lyon 1, F-69000 Lyon, France; Réseau NETSARC +, F69000 Lyon, France; Equipe EMS, Centre Léon Bérard, F-69000 Lyon, France; Département d’Oncologie Médicale, Institut Bergonié, F-33000 Bordeaux, France; ULR 2694 - Metrics: Evaluation des technologies de santé et des pratiques médicales, CHU de Lille, Univ. Lille, F-59000 Lille, France; Département de Biopathologie, Institut Bergonié, F-33000 Bordeaux, France; Department of Pathology, Trousseau University Hospital, F-37000 Tours, France; Unité de recherche clinique et épidémiologique, Inserm CIC 1401, Institut Bergonié, F-33000 Bordeaux

## Abstract

**Objective:** Sarcomas are rare tumors of connective tissue that can develop in soft-tissue, viscera organs, or bones. Previous occupational studies have mainly focused on men and soft-tissue sarcomas. This study describes associations between occupations and sarcomas, in men and women, for soft-tissue sarcomas (STS), including visceral sarcomas, and bone sarcomas (BS).

**Methods:** The ETIOSARC study was a multicenter case-control study conducted across six French geographical areas between 2019 and 2023. Occupational histories were collected by interview and coded according to the International Standard Classification of Occupations (2008). Conditional logistic regression models were applied to estimate odds ratios with 90% confidence intervals for each occupation included in the study.

**Results:** A total of 374 male cases (336 STS and 38 BS), 371 female cases (335 STS and 36 BS), and 1,387 controls were included. Positive associations were observed among male STS for painters (OR=5.37, 90% CI=1.83–15.71), waiters (OR=4.54, 90% CI=1.88–10.97), and among male BS for electricians (OR=3.67, 90% CI=0.91–14.86). Among women, increased STS risk was found among real estate agents (OR=3.48, 90% CI=1.03–11.79), food preparation assistants (OR=3.47, 90% CI=1.09–11.04), and administrative and executive secretaries (OR=2.37, 90% CI=1.45–3.86). For BS, a higher risk was observed among sales workers (OR=5.61, 90% CI=1.65–18.99).

**Conclusions:** Our study highlights hitherto unreported occupational associations with sarcomas, among both men and women, as well as the importance of sex-stratified analyses. Further research should aim to confirm these results and disentangle the respective roles of occupational exposures, environmental factors, and lifestyle characteristics.

## Introduction

Sarcomas are rare, heterogeneous connective tissue tumors that can arise at any anatomical site. More than 150 histological subtypes have been identified (1). Although sarcomas are among the most common cancers in children, they represent less than 1% of adult cancers (2). Three main groups of sarcomas can be identified: soft-tissue sarcomas (STS), visceral sarcomas (VS, including gastrointestinal stromal tumors [GIST]), and bone sarcomas (BS). STS and VS are the most frequent and together account for more than 80% of sarcomas. Diagnosing a sarcoma requires expert pathological review to avoid misclassification between histological types or with non-mesenchymal mimickers (3). The wide variety of tumors, the complexity of diagnosis, and the rarity of the disease mean that the incidence of sarcomas is incompletely documented, and their etiology is little studied and often contradictory. In recent years, the incidence of all sarcomas appears to have increased in both men and women, with variations according to primary site and histological subtypes (4–6). While STS incidence has stabilized, BS and GIST incidences continue to rise. Across all sarcomas, the sex ratio (M/F) is close to 1, but large variations are observed by histological type, ranging from 0.5 for angiosarcoma to 1.8 for liposarcoma (5). Nonetheless, incidence is higher overall among men than among women (5,7). This increase in incidence, along with differences according to sex and histological type, suggests that environmental factors may be implicated in their etiology.

Only a few studies have explored the etiology of sarcomas, and those that exist have focused almost exclusively on men and STS. The only established risk factors are ionizing radiation (including radiotherapy) and heritable syndromes (2,8). Suspected factors include phenoxy-herbicides, chlorophenols, and wood dust for STS (9–11), as well as vinyl chloride for liver angiosarcomas (12). Occupations exposed to these factors have been associated with sarcomas in some studies: farmers (13), gardeners (14,15), and construction workers among male STS (14); cattle farmers among men and women for all sarcomas (16); and woodworkers among male and female BS (17,18). But other studies have reported no associations for these same occupations including farmers among STS and BS (14,17,19) or construction workers (17). Moreover, additional occupations have been identified, without a clear established causal agent including machinists (20) and firefighters (21) among male STS, and blacksmiths, toolmakers, and machine tool operators among male and female BS (17). Except for this latest case-control study on BS, which included both men and women, all these studies examined associations between occupations and sarcomas among men only. The only study conducted exclusively among women is an occupational cohort investigating the link between occupations and uterine leiomyosarcoma and endometrial stromal sarcoma. Associations were found for farmers, teachers, and shoe and leather workers in relation to uterine leiomyosarcoma, but not for gardeners, woodworkers, or other construction workers (22).

Taking sex into account when studying the occupational etiology of sarcomas seems essential. First, differences in the distribution of histological sarcoma subtypes between men and women, with specific tumors affecting female reproductive organs, may reflect distinct mechanisms underlying carcinogenesis in response to occupational factors (23). Second, occupational exposure differs depending on sex (24). Men are more often exposed to dust and chemical agents and use more vibrating tools, whereas women report performing repetitive tasks or working at very high speeds. These differences remain even within the same occupation (25). In addition, it is important to consider primary site when studying sarcoma etiology. Differences in incidence patterns by sex and site raise the hypothesis of distinct carcinogenic mechanisms and, therefore, distinct etiologies.

Previous studies on the role of occupations in sarcomas have been inconsistent; most have been conducted only with relation to men, almost exclusively for STS, and had small sample sizes. The objective of our research is to describe associations between occupations and sarcomas separately for men and women, distinguishing soft tissue sarcomas (including visceral sarcomas) from bone sarcomas. To address this objective, we have used data from the French ETIOSARC study, the largest occupational case-control study on sarcomas to date.

## Methods

### Study population

The ETIOSARC study protocol has been previously described (26). Population-based cancer registries and the French network of sarcoma experts, NETSARC +, identified cases in five geographic areas (Gironde, Lille, Haut-Rhin, Isère, and Poitou-Charentes), whereas only NETSARC+ identified cases in the Rhône department (1). All cases were newly diagnosed primary sarcoma patients, including patients with soft tissue (STS), visceral (VS), and bone sarcomas (BS), according to the WHO classification of tumors of soft tissue and bone (4th edition, 2013) (27). Diagnoses were made between February 2019 and March 2023. At the time of diagnosis, the patients were aged between 18 to 80 and resided in one of the participating areas. Cases with Kaposi sarcomas were excluded, as were those with known genetic syndromes linked to an increased risk of developing sarcomas, including Li-Fraumeni syndrome, retinoblastoma syndrome, neurofibromatosis, and Ollier disease. Histological confirmation of sarcoma diagnosis was obtained through centralized expert review by NETSARC+ pathologists. Shortly after case identification and prior to patient contact, general practitioners were contacted to obtain information on patients’ health conditions.

For each case included, two controls were randomly selected from electoral rolls. Controls were individually matched according to sex, age (in 5-year age groups), and geographic area. To ensure the representativeness of controls, the selection process was carefully documented in the protocol (26).

### Data collection

Cases and controls were interviewed using an identical procedure. After obtaining oral consent, participants received a self-administered questionnaire by mail. The questionnaire collected complete residential and occupational histories, as well as occupational information for spouses and parents. For each job held for more than six months, information such as start and end dates, company name, sector of activity, occupation, and duration of employment was recorded. Subsequently, a trained clinical research assistant conducted a face-to-face interview after participants had signed a consent form. The occupational calendar was completed with detailed descriptions of each job, including tasks performed, co-activities, materials, products, and tools used. A second questionnaire collected information on demographic and socioeconomic characteristics, occupational and domestic exposures, leisure activities, lifestyle factors (tobacco use, alcohol consumption, and diet), medical history, family history of cancer, and reproductive history for women. At the end of the interview, participants who consented provided a saliva sample. Between March 2020 and May 2021, due to the COVID-19 pandemic, interviews were conducted by phone or video conference. In 2022, proxy interviews were conducted for cases who died shortly after diagnosis. These shorter questionnaires focused mainly on occupational history and exposures. Controls matched to deceased cases were interviewed with these shorter questionnaires.

### Coding of occupations

All occupations listed in the questionnaires were coded by trained research assistants using the International Standard Classification of Occupations (ISCO-08). A subject was considered exposed if they had worked in a given occupation for at least 6 months, based on the duration reported by the subject, without latency period. Exposure was then categorized as: never exposed, 6 months to 5 years, and more than 5 years.

### Statistical analyses

For each ISCO sub-major group (2-digit codes) with at least 5 exposed cases, conditional logistic regression models were used to estimate odds ratios (OR) with 90% confidence intervals (90% CI) to avoid overly restrictive interpretation of the results (28). For occupations showing a significant excess risk, ORs and 90% CIs were further estimated at a finer level (unit groups at 4 digits) where at least 5 cases were available. Duration analyses were conducted for occupations with OR exceeding 1.5. In these analyses, the 5-case threshold was not applied. The reference group for each model consisted of subjects who had never worked in the occupation in question or had done so for less than 6 months. A specific grouping for agricultural occupations was created, combining ISCO major group 6 (skilled agricultural, forestry, and fishery workers) and sub-major group 92 (agricultural, forestry, and fishery laborers). Similarly, woodworkers were grouped into a single category, including minor group 752 (wood treaters, cabinet-makers and related trades workers), unit groups 7115 (carpenters and joiners), 7317 (handicraft workers in wood, basketry and related materials), and 8172 (wood processing plant operators).

Confounding factors were identified by constructing a directed acyclic graph (DAG) in which exposure was defined as lifetime occupational history and the outcome as all sarcomas (Supplementary Material Figure S1) (29). Given the limited number of established sarcoma risk factors, potential confounding was considered in a broader cancer epidemiology context. Education level was identified as the only relevant confounder and was considered as a proxy for socioeconomic status. It was categorized into 3 groups (secondary, 1-to 2-year university degree, and ≥3-year university degree). Age was included in all models, as a continuous variable, even though controls were matched by age, to account for residual variability within the sets.

Cases or controls who had never worked or had worked for less than 6 months, were excluded from the analyses as well as their matched controls or cases.

All analyses were performed separately for men and women for STS (including VS) and BS using RStudio 2026.01.2.

We conducted sensitivity analyses restricted to histologically confirmed cases and their matched controls.

## Results

### Characteristics of the study population

Of the 1,497 cases identified, 1,257 were contacted to participate, and 770 were included in the ETIOSARC study (Supplementary Material Figure S2). For controls, 13,924 subjects were randomly selected; 4,820 were contacted, and 1,441 were included. The response rates were 61.3% for cases and 29.9% for controls. After exclusions (n=25 cases), 745 were included in the analyses: 671 STS (336 men and 335 women) and 74 BS (38 men and 36 women). A total of 1,387 controls were included in the final analyses. Proxy interviews were conducted for 11 STS cases (19 controls) and 1 BS case (2 controls) only.

Main characteristics of cases and controls are shown in Table 1, categorized by tumor site and sex. Men and women were equally represented. The median age was 60.4 years for male STS cases and 59.2 years for female STS cases, while for BS it was 53.5 years in men and 57.6 years in women. Controls had higher education levels than cases, regardless of sex or tumor site: between 44.4% to 47.1% of controls had at least a 3-year university degree versus 27.8% to 40.6% for cases. On average, men reported a higher number of jobs than women (5 versus 4) and a longer working lifetime (between 30 to 35 years in men versus 26 to 28 years in women). Socio-professional category, based on the most recent occupation, differed between cases and controls. A higher proportion of craft and related trades workers was observed among male cases compared with controls (14.9% among STS and 13.2% among BS cases versus 8.6% and 1.6% in control groups, respectively). Among women, controls were more frequently classified as professionals (34.1% and 35.3% in the control groups) than cases (26.6% for STS and 19.4% for BS).

**Table 1:**
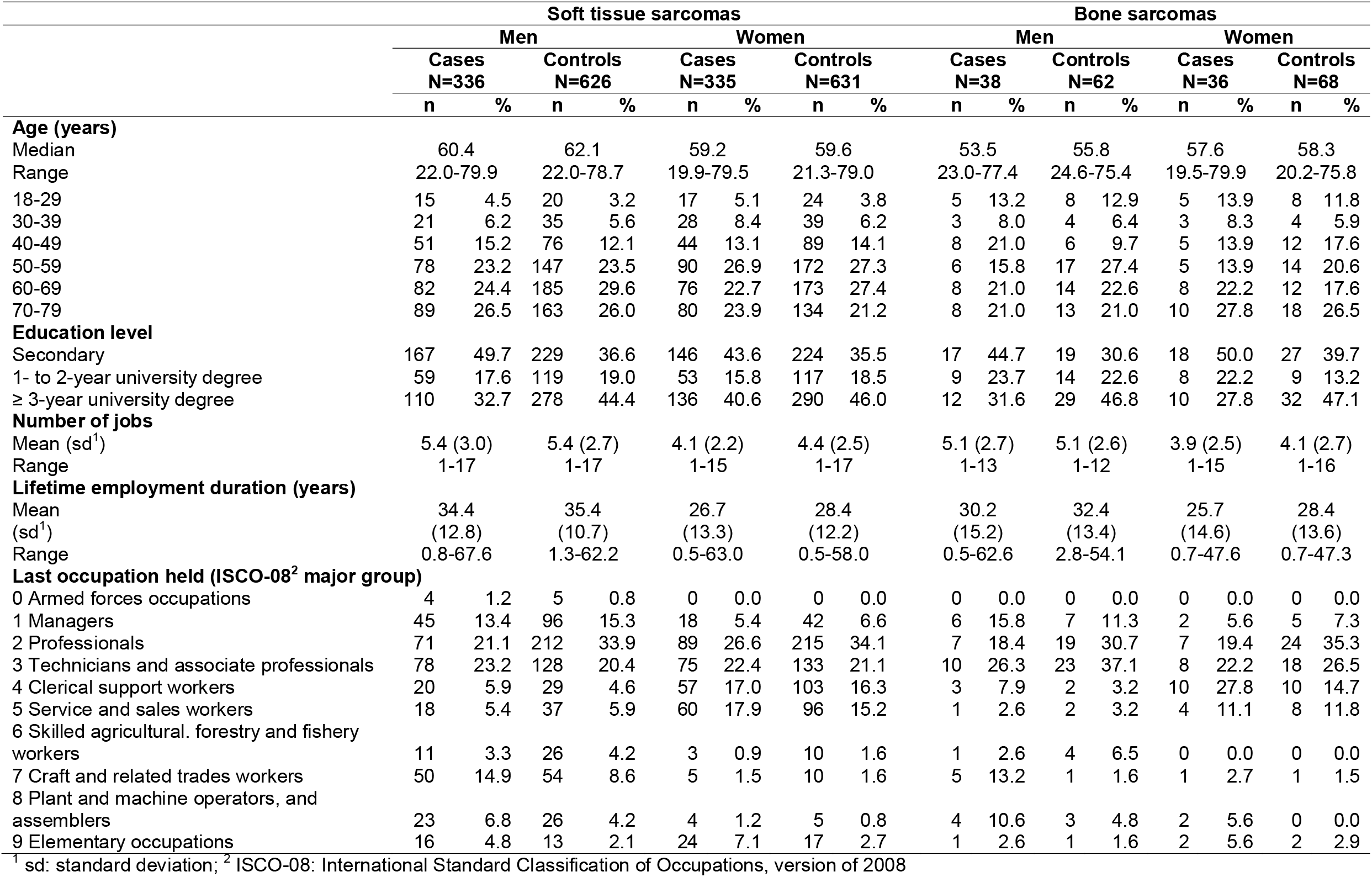
Main characteristics of sarcoma cases (N=745) depending on the location and gender, and controls (N=1,387) – ETIOSARC study, France, 2019-2023.

|  | Soft tissue sarcomas |  |  |  |  |  |  |  | Bone sarcomas |  |  |  |  |  |  |  |
| --- | --- | --- | --- | --- | --- | --- | --- | --- | --- | --- | --- | --- | --- | --- | --- | --- |
|  | Men |  |  |  | Women |  |  |  | Men |  |  |  | Women |  |  |  |
|  | Cases<br>N=336 |  | Controls<br>N=626 |  | Cases<br>N=335 |  | Controls<br>N=631 |  | Cases<br>N=38 |  | Controls<br>N=62 |  | Cases<br>N=36 |  | Controls<br>N=68 |  |
|  | n | % | n | % | n | % | n | % | n | % | n | % | n | % | n | % |
| **Age (years)** |  |  |  |  |  |  |  |  |  |  |  |  |  |  |  |  |
| Median | 60.4 |  | 62.1 |  | 59.2 |  | 59.6 |  | 53.5 |  | 55.8 |  | 57.6 |  | 58.3 |  |
| Range | 22.0-79.9 |  | 22.0-78.7 |  | 19.9-79.5 |  | 21.3-79.0 |  | 23.0-77.4 |  | 24.6-75.4 |  | 19.5-79.9 |  | 20.2-75.8 |  |
| 18-29 | 15 | 4.5 | 20 | 3.2 | 17 | 5.1 | 24 | 3.8 | 5 | 13.2 | 8 | 12.9 | 5 | 13.9 | 8 | 11.8 |
| 30-39 | 21 | 6.2 | 35 | 5.6 | 28 | 8.4 | 39 | 6.2 | 3 | 8.0 | 4 | 6.4 | 3 | 8.3 | 4 | 5.9 |
| 40-49 | 51 | 15.2 | 76 | 12.1 | 44 | 13.1 | 89 | 14.1 | 8 | 21.0 | 6 | 9.7 | 5 | 13.9 | 12 | 17.6 |
| 50-59 | 78 | 23.2 | 147 | 23.5 | 90 | 26.9 | 172 | 27.3 | 6 | 15.8 | 17 | 27.4 | 5 | 13.9 | 14 | 20.6 |
| 60-69 | 82 | 24.4 | 185 | 29.6 | 76 | 22.7 | 173 | 27.4 | 8 | 21.0 | 14 | 22.6 | 8 | 22.2 | 12 | 17.6 |
| 70-79 | 89 | 26.5 | 163 | 26.0 | 80 | 23.9 | 134 | 21.2 | 8 | 21.0 | 13 | 21.0 | 10 | 27.8 | 18 | 26.5 |
| **Education level** |  |  |  |  |  |  |  |  |  |  |  |  |  |  |  |  |
| Secondary | 167 | 49.7 | 229 | 36.6 | 146 | 43.6 | 224 | 35.5 | 17 | 44.7 | 19 | 30.6 | 18 | 50.0 | 27 | 39.7 |
| 1- to 2-year university degree | 59 | 17.6 | 119 | 19.0 | 53 | 15.8 | 117 | 18.5 | 9 | 23.7 | 14 | 22.6 | 8 | 22.2 | 9 | 13.2 |
| ≥ 3-year university degree | 110 | 32.7 | 278 | 44.4 | 136 | 40.6 | 290 | 46.0 | 12 | 31.6 | 29 | 46.8 | 10 | 27.8 | 32 | 47.1 |
| **Number of jobs** |  |  |  |  |  |  |  |  |  |  |  |  |  |  |  |  |
| Mean (sd<sup>1</sup>) | 5.4 (3.0) |  | 5.4 (2.7) |  | 4.1 (2.2) |  | 4.4 (2.5) |  | 5.1 (2.7) |  | 5.1 (2.6) |  | 3.9 (2.5) |  | 4.1 (2.7) |  |
| Range | 1-17 |  | 1-17 |  | 1-15 |  | 1-17 |  | 1-13 |  | 1-12 |  | 1-15 |  | 1-16 |  |
| **Lifetime employment duration (years)** |  |  |  |  |  |  |  |  |  |  |  |  |  |  |  |  |
| Mean (sd<sup>1</sup>) | 34.4 (12.8) |  | 35.4 (10.7) |  | 26.7 (13.3) |  | 28.4 (12.2) |  | 30.2 (15.2) |  | 32.4 (13.4) |  | 25.7 (14.6) |  | 28.4 (13.6) |  |
| Range | 0.8-67.6 |  | 1.3-62.2 |  | 0.5-63.0 |  | 0.5-58.0 |  | 0.5-62.6 |  | 2.8-54.1 |  | 0.7-47.6 |  | 0.7-47.3 |  |
| **Last occupation held (ISCO-08<sup>2</sup> major group)** |  |  |  |  |  |  |  |  |  |  |  |  |  |  |  |  |
| 0 Armed forces occupations | 4 | 1.2 | 5 | 0.8 | 0 | 0.0 | 0 | 0.0 | 0 | 0.0 | 0 | 0.0 | 0 | 0.0 | 0 | 0.0 |
| 1 Managers | 45 | 13.4 | 96 | 15.3 | 18 | 5.4 | 42 | 6.6 | 6 | 15.8 | 7 | 11.3 | 2 | 5.6 | 5 | 7.3 |
| 2 Professionals | 71 | 21.1 | 212 | 33.9 | 89 | 26.6 | 215 | 34.1 | 7 | 18.4 | 19 | 30.7 | 7 | 19.4 | 24 | 35.3 |
| 3 Technicians and associate professionals | 78 | 23.2 | 128 | 20.4 | 75 | 22.4 | 133 | 21.1 | 10 | 26.3 | 23 | 37.1 | 8 | 22.2 | 18 | 26.5 |
| 4 Clerical support workers | 20 | 5.9 | 29 | 4.6 | 57 | 17.0 | 103 | 16.3 | 3 | 7.9 | 2 | 3.2 | 10 | 27.8 | 10 | 14.7 |
| 5 Service and sales workers | 18 | 5.4 | 37 | 5.9 | 60 | 17.9 | 96 | 15.2 | 1 | 2.6 | 2 | 3.2 | 4 | 11.1 | 8 | 11.8 |
| 6 Skilled agricultural, forestry and fishery workers | 11 | 3.3 | 26 | 4.2 | 3 | 0.9 | 10 | 1.6 | 1 | 2.6 | 4 | 6.5 | 0 | 0.0 | 0 | 0.0 |
| 7 Craft and related trades workers | 50 | 14.9 | 54 | 8.6 | 5 | 1.5 | 10 | 1.6 | 5 | 13.2 | 1 | 1.6 | 1 | 2.7 | 1 | 1.5 |
| 8 Plant and machine operators, and assemblers | 23 | 6.8 | 26 | 4.2 | 4 | 1.2 | 5 | 0.8 | 4 | 10.6 | 3 | 4.8 | 2 | 5.6 | 0 | 0.0 |
| 9 Elementary occupations | 16 | 4.8 | 13 | 2.1 | 24 | 7.1 | 17 | 2.7 | 1 | 2.6 | 1 | 1.6 | 2 | 5.6 | 2 | 2.9 |

<sup>1</sup> sd: standard deviation; <sup>2</sup> ISCO-08: International Standard Classification of Occupations, version of 2008
<sup>1</sup> ISCO-08 International Standard Classification of Occupations, version of 2008

Histological confirmation was obtained for the vast majority of cases: 93.8% in men and 90.4% in women with STS, and 97.4% in men and 83.8% in women with BS (Supplementary Material Table S1). Among men with STS, the most frequent histological subtypes were liposarcoma (25.3%) and unclassified and undifferentiated sarcomas (14.9%). Among women, leiomyosarcoma (20.0%) and liposarcoma (16.1%) were the most common. Among both men and women with BS, chondrosarcomas (34.2% in men and 52.7% in women) and chordomas (26.3% and 19.4% respectively) were the most frequent histological subtypes.

### Occupations and Soft Tissue Sarcomas

Positive associations with STS were observed among men (Figure 1) for non-commissioned armed forces officers (ISCO 02), personal service workers (ISCO 51), building and related trades workers (ISCO 71), and refuse workers and other elementary workers (ISCO 96). Drivers and mobile plant operators (ISCO 83) and cleaners and helpers (ISCO 91) tended to be associated with STS. At a more detailed level, waiters (ISCO 5131) (OR=4.54, 90% CI=1.88–10.97) and painters and related workers (ISCO 7131) (OR=5.37, 90% CI=1.83–15.71) were associated with STS. Conversely, negative associations were observed for business and administration professionals (ISCO 24) (OR=0.57, 90% CI=0.37–0.89) and other clerical support workers (ISCO 44) (OR=0.40, 90% CI=0.19–0.86).

**Figure 1:**
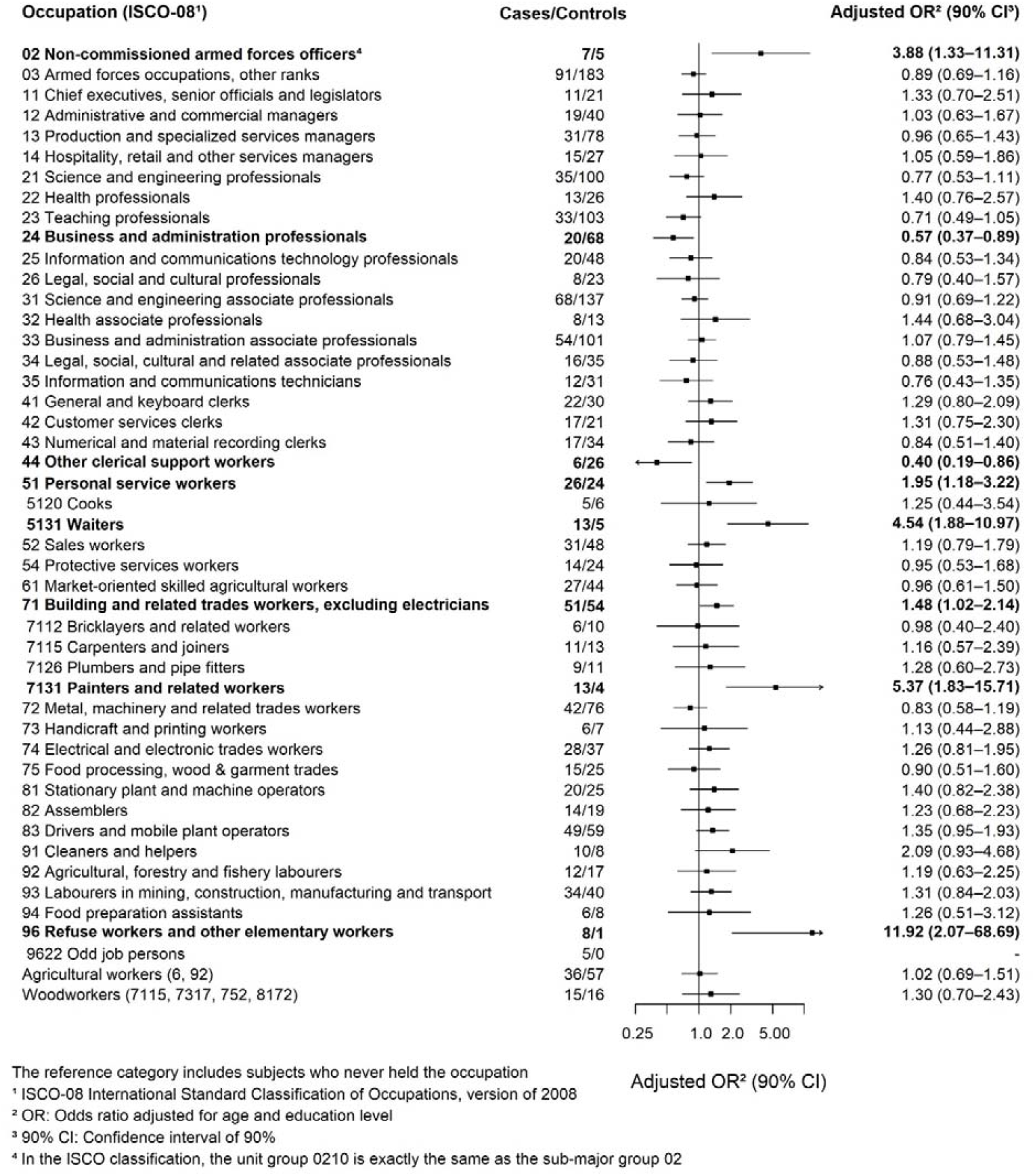
Odds ratios and 90% confidence intervals (adjusted for age and education level) for the association between occupations (with at least 5 cases) and soft tissue sarcomas among men (336 cases / 626 controls) – ETIOSARC study, France, 2019-2023

Among women, occupations associated with STS included business and administration associate professionals (ISCO 33) and food preparation assistants (ISCO 94), whereas cleaners and helpers (ISCO 91) were likely to be associated (Figure 2). More specifically, occupations associated with STS were real estate agents and property managers (ISCO 3334) (OR=3.48, 90% CI=1.03–11.79), administrative and executive secretaries (ISCO 3343) (OR=2.37, 90% CI=1.45–3.86), and kitchen helpers (ISCO 9412) (OR=2.99, 90% CI=0.91–9.85).

**Figure 2:**
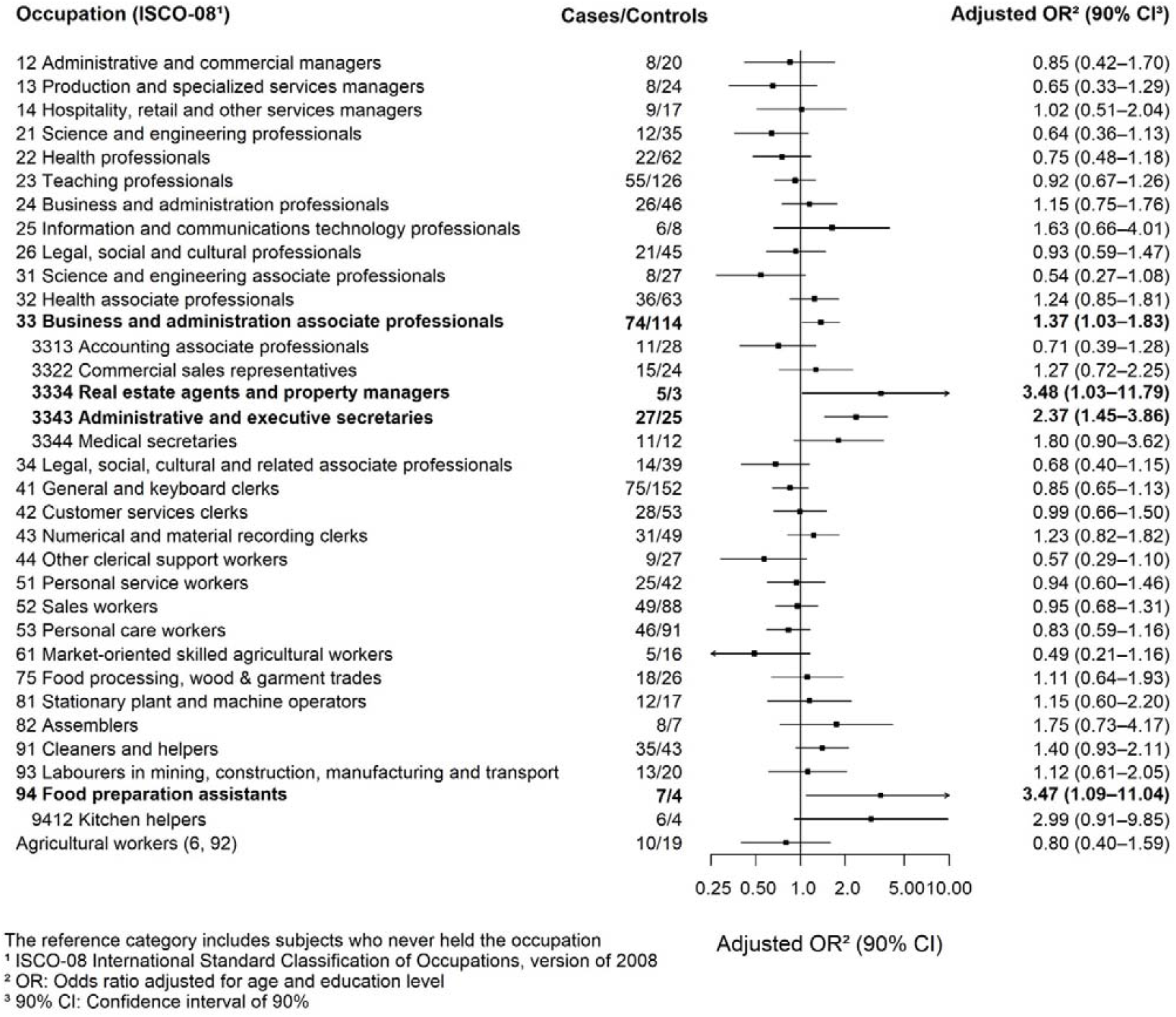
Odds ratios and 90% confidence intervals (adjusted for age and education level) for the association between occupations (with at least 5 cases) and soft tissue sarcomas among women (335 cases and 631 controls) – ETIOSARC study, France, 2019-2023

No association has been identified between agricultural workers and STS in men (OR=1.02, 90% CI=0.69–1.51). Among women, the relationship appeared negative (OR=0.80, 90% CI=0.40–1.59). The OR for woodworkers was above 1 in men, suggesting a positive association (OR=1.30, 90% CI=0.70–2.43).

Results are similar even without adjusting for education level (Supplementary Materials, Figures S3–S4).

Duration-based analyses showed a higher risk among men for non-commissioned armed forces officers (ISCO 02) and painters and related workers (ISCO 7131) where occupations were held for 5 years or more (Supplementary Materials Table S2). Among women, a positive association was observed for administrative and executive secretaries (ISCO 3343).

Sensitivity analyses restricted to histologically confirmed cases showed largely similar results, but for occupations with a small number of cases, some estimates differed slightly, although overall patterns remained consistent (Supplementary Materials, Figures S5–S6).

### Occupations and Bone Sarcomas

Among men, electrical and electronic trades workers (ISCO 74) (OR=3.67, 90% CI=0.91–14.86) tended to be associated with BS (Figure 3). Among women, sales workers (ISCO 52) (OR=5.61, 90% CI=1.65–18.99) were associated with BS (Figure 4).

**Figure 3:**
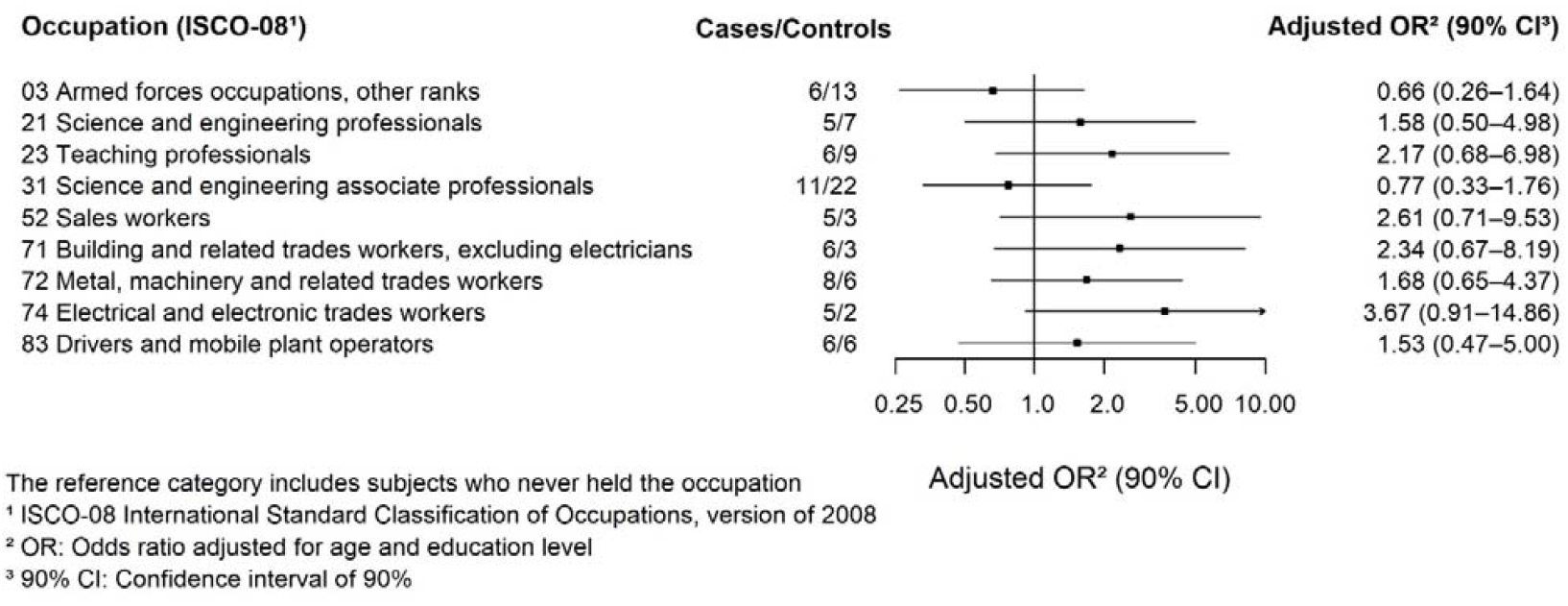
Odds ratios and 90% confidence intervals (adjusted for age and education level) for the association between occupations (with at least 5 cases) and bone sarcomas among men (38 cases / 62 controls) – ETIOSARC study, France, 2019-2023

**Figure 4:**
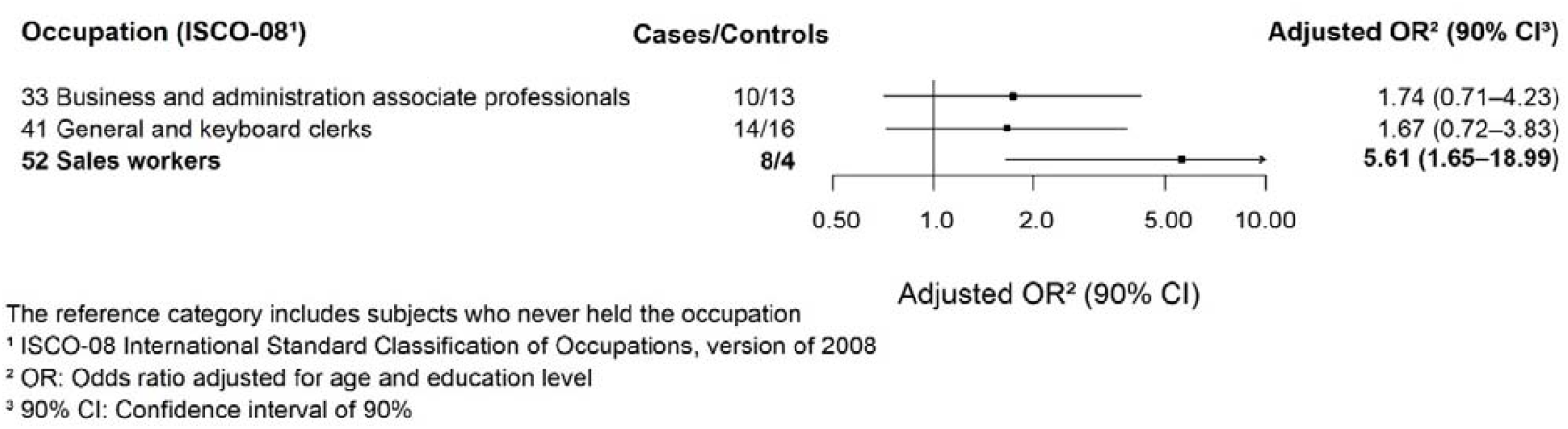
Odds ratios and 90% confidence intervals (adjusted for age and education level) for the association between occupations (with at least 5 cases) and bone sarcomas among women (36 cases and 68 controls) – ETIOSARC study, France, 2019-202

Agricultural workers and woodworkers accounted for only 2 male cases each, and no female cases.

As with STS, results are similar even without adjusting for education level (Supplementary Materials, Figures S7–S8).

Analysis by duration of employment did not show higher risk when time spent in these occupations was above 5 years (Supplementary Materials, Table S3).

Analyses restricted to histologically confirmed cases resulted in reduced sample sizes, but findings were broadly consistent and pointed in the same direction (Supplementary Material, Figures S9– S10).

## Discussion

This study investigated associations between occupations and the occurrence of sarcomas by tumor site and by sex in a large population-based case-control study. To our knowledge, it is the first case-control study with such a large number of cases to detail the occupational risks of sarcomas for women. Among men, associations were found for non-commissioned armed forces officers, waiters, and painters for STS, and for electricians for BS. Among women, real estate agents and property managers, administrative and executive secretaries, and food preparation assistants were associated with STS, while sales workers were associated with BS.

Only a limited number of studies have investigated occupational risks of sarcomas, often with outdated data and different classifications, making comparisons difficult. However, some of our results are consistent with previous studies. Military occupations are been associated with STS in one study (30) but not in another (20). When grouping all military ranks (ISCO major group 0), we found no association (OR=0.97, 90% CI = 0.71–1.31), suggesting that our more detailed coding may explain the association identified in our study for non-commissioned armed forces officers. Construction-related occupations have already been associated with STS: Wingren *et al*. (14) reported an excess risk among workers exposed to asbestos and other insulating materials. In our study, associations with STS were found in several sub-major groups: building and related trades workers (ISCO 71); labourers in mining, construction, manufacturing and transport (ISCO 93); and refuse workers and other elementary workers (ISCO 96), where 5 out 8 cases were employed as odd-job persons (ISCO 9622) who performed maintenance tasks. Electricians (ISCO 74), some involved in construction, were associated with BS in men. These occupations involve exposure to multiple carcinogens (e.g., asbestos, silica, wood dust). Previous study have also reported associations for cleaners (ISCO 91) and drivers (ISCO 83) (30).

We identified excess risks in occupations that have not hitherto been reported: waiters and painters for male STS; real estate agents and property managers, administrative and executive secretaries, and food preparation assistants for female STS; and sales workers for female BS. Increased cancer risk among waiters may relate to tobacco and alcohol exposure (31–33), although their role in sarcoma remains unclear. Although not specifically linked to sarcoma, painters are exposed to multiple carcinogens (solvents, formaldehyde, cadmium, benzene, …) and are classified by the IARC as Group 1 (34). For the occupations identified in women, to our knowledge, no prior associations with cancer have been reported, warranting further investigations into sector-specific exposures.

We did not observe any association between agricultural work and sarcomas, despite previously reported links with phenoxy herbicides and chlorophenols (10,17,19). Other studies also reported no association with farming (11,19,20). As our approach does not directly assess exposures, targeted analyses are needed.

### Strengths and limitations

Our study is one of the few to examine sex-specific occupational cancer risks. Our findings support the hypothesis of sex differences in occupational exposures, suggesting distinct mechanisms underlying the development of cancer cells in response to exposure factors. Indeed, some associations differed by sex: whereas business and administration associate professionals (ISCO 33) were associated with STS among women, the OR was close to 1 among men; conversely, an excess risk of STS was found for male personal service workers (ISCO 51), while no association was found for women.

Stratification by sex was feasible because the sarcoma sample size in our study is among the largest in occupational case-control studies. For STS, only Hossain *et al*. (20) reported a comparable number of cases (357 male cases), although only 58.9% were histologically confirmed. In our study, more than 90% of cases were histologically confirmed. Sensitivity analyses restricted to histologically confirmed cases yielded similar results. We identified only one study reporting associations between occupations and BS, based on 96 cases, including both men and women (17). Our study, therefore, adds more recent data to the limited existing literature.

Although our study is the largest occupational case-control study of sarcomas to date, the sample size did not allow analysis at the most detailed level. This may have limited our ability to detect some associations due to insufficient statistical power and to explore risks across specific histological subtypes. In addition, given the number of tests performed, some findings may be due to chance and should be interpreted with caution.

Case identification through population-based cancer registries and the NETSARC+ network enabled us to ensure high completeness of case identification within the study area, thereby reducing selection bias. To ensure the representativeness of the controls, we selected them from the electoral rolls, which cover nearly 90% of the French adult population. Our selection process aimed to contact as many eligible controls as possible without introducing selection. Participation rates (61% among cases, and 30% among controls) were lower than the median reported in earlier occupational case-control studies (35), although such rates have declined over time. As exposure data were unavailable for non-participants, we compared included and non-included individuals using the European Deprivation Index (EDI) based on residential addresses at enrollment (36). Included participants were slightly less deprived (24.8% of included cases in quintile 1 *vs*. 19.7% of non-included and 28.3% and 23.2% for controls, respectively)—a common pattern in epidemiological studies (37). However, as these differences were similar among cases and controls, and as occupational exposures concern both white- and blue-collar workers, selection bias is likely to be limited. To further account for socioeconomic differences, we decided to adjust for education level (38).

Recall bias is unlikely to have substantially affected our results, as no well-established occupational risk factor for sarcomas exists, and cases had no clear reason to recall their occupational history differently from controls. Retrospective collection of complete occupational histories may have introduced some misclassification, but such errors are expected to be non-differential.

Among potential confounders, only education level was retained in the models. Although confounding is ideally assessed for each exposure-disease pair using specific DAGs, we constructed a single one for simplicity, considering all occupations as exposure and all sarcomas as the outcome. Smoking, alcohol consumption, BMI, and ionizing radiation exposure were not included as covariates, as they were considered potential mediators rather than confounders.

## Conclusion

In conclusion, our study provides new insights into the occupational etiology of sarcomas, identifying occupations never previously associated with STS or BS among men, such as waiters, painters, and electricians. However, it does not confirm previously reported associations with farmers and woodworkers. The unexpected results in women should prompt researchers to conduct focused studies on women. These findings represent an initial step toward a better understanding on the occupational determinants of sarcomas. Further analyses are needed, to investigate specific exposures such as pesticides. Finally, both broadly defined environmental exposures and lifestyle factors should be investigated.

## Data Availability

All data produced in the present study are available upon reasonable request to the authors

## Ethic statement

This study is part of clinical trial C17-03 sponsored by Inserm. It was granted approval by local Ethics Committee or “Comité de Protection des Personnes” on 05/16/2018, authorized by the French authorities and registered in a public trials registry (CT NCT03670927).

All study participants gave their informed, written consent to participation, in line with French ethical guidelines.

## Data available statement

Data are available upon reasonable request

## Acknowledgements

The authors would like to thank all participants (cases, next of kin, and controls) for their valuable contribution to this study.

The authors are grateful to the NETSARC+ and INTERSARC networks for their contribution to this study.

The authors also thank all the clinical research associates, data managers, and center managers for their essential work in data collection, conducting participant interviews, coding, and data entry for questionnaires and data management.

EPICENE team: L. Ayayi, A. Barhoumi, M. Barault, S. Constant, M. Depont, J. Gallet, N. Kabbani,

P. Koenig, N. Lefebvre, J. Orizono, I. Perrot, S. Rasanda, A. Tedlaouti

Bergonié Institut: JB. Courrèges, A. Giraud, N. Huchet

Gironde cancer registry : C. Galy, M. Rabet, E. Tranchet Isère cancer registry: C. Camara

Lille cancer registry: V. Démarret, S. Plouvier

Haut-Rhin cancer registry: V. Beck, L. Sasselli

GHRMSA Clinical Research Unit (Haut-Rhin): J. Bontemps, E. Bouchet, E. Cormier, S. Ledy Léon Bérard Center (Rhône and Isère): N. Akrim-Durand, M. Girodet, C. Maqua

Poitou-Charentes registry: G. Defossez, C. Dudognon, V. Martin, N. Mériau

