## Supplementary materials for "Associations between occupations and the occurrence of sarcomas: results of the French population-based case-control study ETIOSARC"

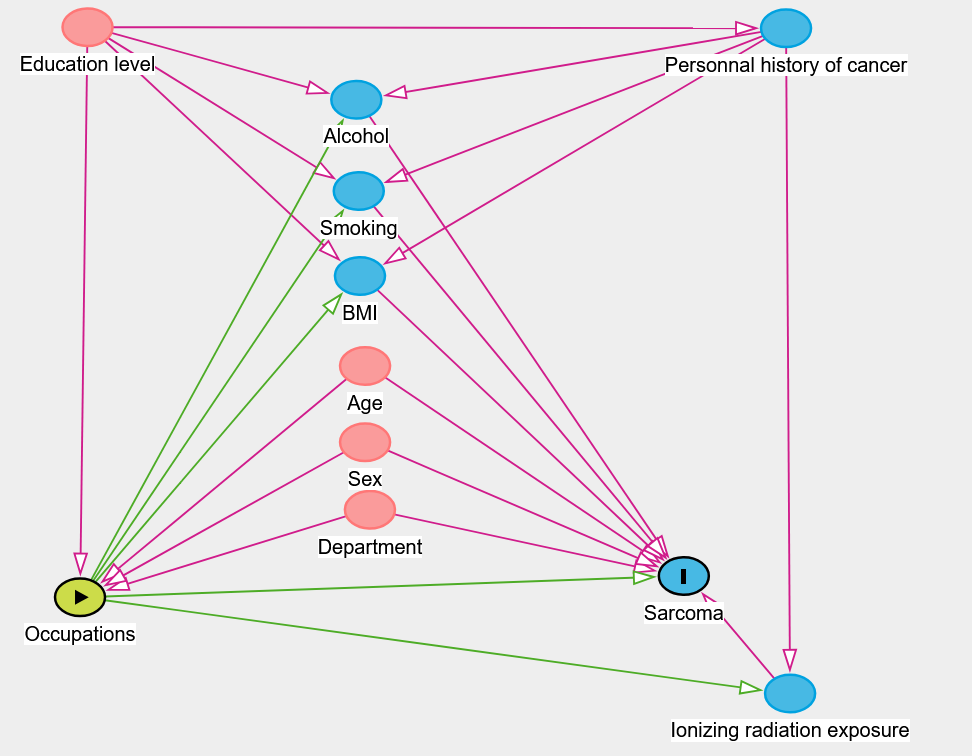

| 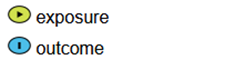 | 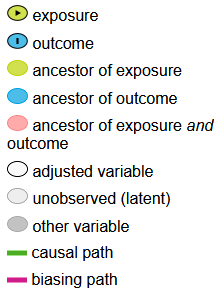 | 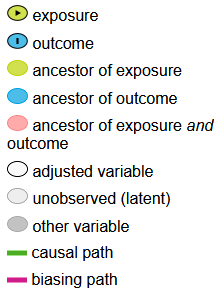 |
| --- | --- | --- |

BMI: body mass index

Figure S1: Directed acyclic graph (DAG) of the association between occupations and sarcoma – ETIOSARC study, France, 2019-2023. DAG created with DAGitty version 3.1 (http://dagitty.net/)

Explanations:

- Age, sex, and department are matching variables.
- Education level determines access to occupations. Moreover, a low education level leads to unhealthy lifestyle habits: higher tobacco and alcohol consumption and higher BMI (1). Indirectly, people with a low education level are more at risk of developing cancer (2).
- Alcohol and smoking are represented by consumption at the index date, like measuring BMI. These three factors are associated with several cancers (3) and sarcomas (4–6). However, occupations may be linked to such behavior: working in stressful, precarious, or physically demanding occupations increases tobacco and alcohol consumption (7,8)
- Personal history of cancer: after a diagnosis of cancer, behavior shifts towards a healthier lifestyle: reduction in tobacco and alcohol consumption (9).
- Ionizing radiation exposure (including radiotherapy) is a risk factor for soft tissue and bone sarcomas (10). Several occupations involve exposure to ionizing radiation (11)

11.	IARC. IARC Monographs on the Evaluation of Carcinogenic Risks to Humans, Vol 100. A review of human carcinogens, part D: radiation. Lyon; 2012.

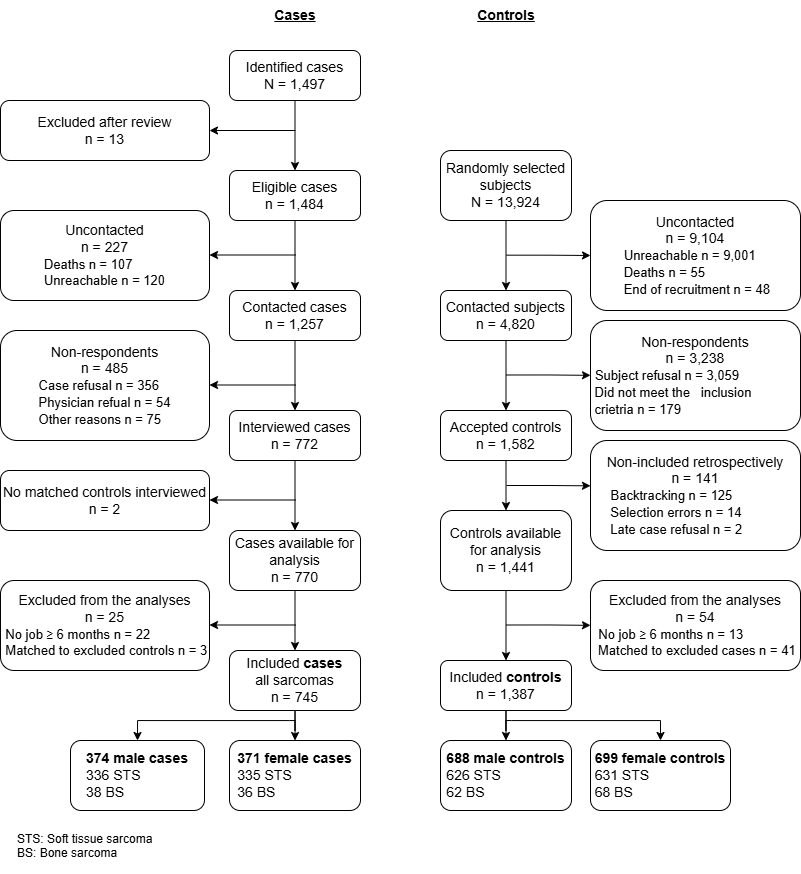

Figure S2: Flow chart of cases and controls included in the study – ETIOSARC study, France, 2019-2023

Table S1: Histological types of sarcoma cases included in the study (N=745) – ETIOSARC study, France, 2019-2023

|  | **Men** | | | **Women** | | |
| --- | --- | --- | --- | --- | --- | --- |
|  | **n** | **%** | **% histological confirmation** | **n** | **%** | **% histological confirmation** |
| **Soft tissue and visceral sarcomas** | **336** | **100.0** | **93.8** | **335** | **100.0** | **90.4** |
| Atypical lipomatous tumor | 43 | 12.8 | 100.0 | 26 | 7.8 | 96.2 |
| GIST (Gastrointestinal stromal tumour) | 35 | 10.4 | 82.9 | 19 | 5.7 | 68.4 |
| Intermediate fibroblastic/myofibroblastic tumors | 27 | 8.0 | 92.6 | 36 | 10.7 | 94.4 |
| Leiomyosarcoma | 27 | 8.0 | 88.9 | 67 | 20.0 | 91.0 |
| Liposarcoma | 85 | 25.3 | 96.5 | 54 | 16.1 | 88.9 |
| Malignant peripheral nerve sheath tumor | 3 | 0.9 | 100.0 | 2 | 0.6 | 100.0 |
| Mesenchymal chondrosarcoma | 0 | 0.0 | - | 1 | 0.3 | 100.0 |
| Miscellaneous sarcomas | 33 | 9.8 | 97.0 | 33 | 9.8 | 93.9 |
| Myxofibrosarcoma | 17 | 5.1 | 94.1 | 15 | 4.5 | 93.3 |
| Other sarcomas | 1 | 0.3 | 100.0 | 28 | 8.4 | 82.1 |
| Rhabdomyosarcoma | 7 | 2.1 | 100.0 | 6 | 1.8 | 83.3 |
| Synovial sarcoma | 8 | 2.4 | 87.5 | 6 | 1.8 | 100.0 |
| Unclassified and undifferentiated sarcomas | 50 | 14.9 | 92.0 | 42 | 12.5 | 97.6 |
| **Bone sarcomas** | **38** | **100.0** | **97.4** | **36** | **100.0** | **83.8** |
| Chondrosarcoma | 13 | 34.2 | 100.0 | 19 | 52.7 | 89.5 |
| Chordomas | 10 | 26.3 | 100.0 | 7 | 19.4 | 71.4 |
| Leiomyosarcoma | 0 | 0.0 | - | 2 | 5.6 | 100.0 |
| Miscellaneous sarcomas | 5 | 13.2 | 100.0 | 2 | 5.6 | 100.0 |
| Osteosarcoma | 7 | 18.4 | 100.0 | 2 | 5.6 | 100.0 |
| Rhabdomyosarcoma | 0 | 0.0 | - | 1 | 2.8 | 100.0 |
| Unclassified and undifferentiated sarcomas | 3 | 7.9 | 66.7 | 3 | 8.3 | 66.7 |

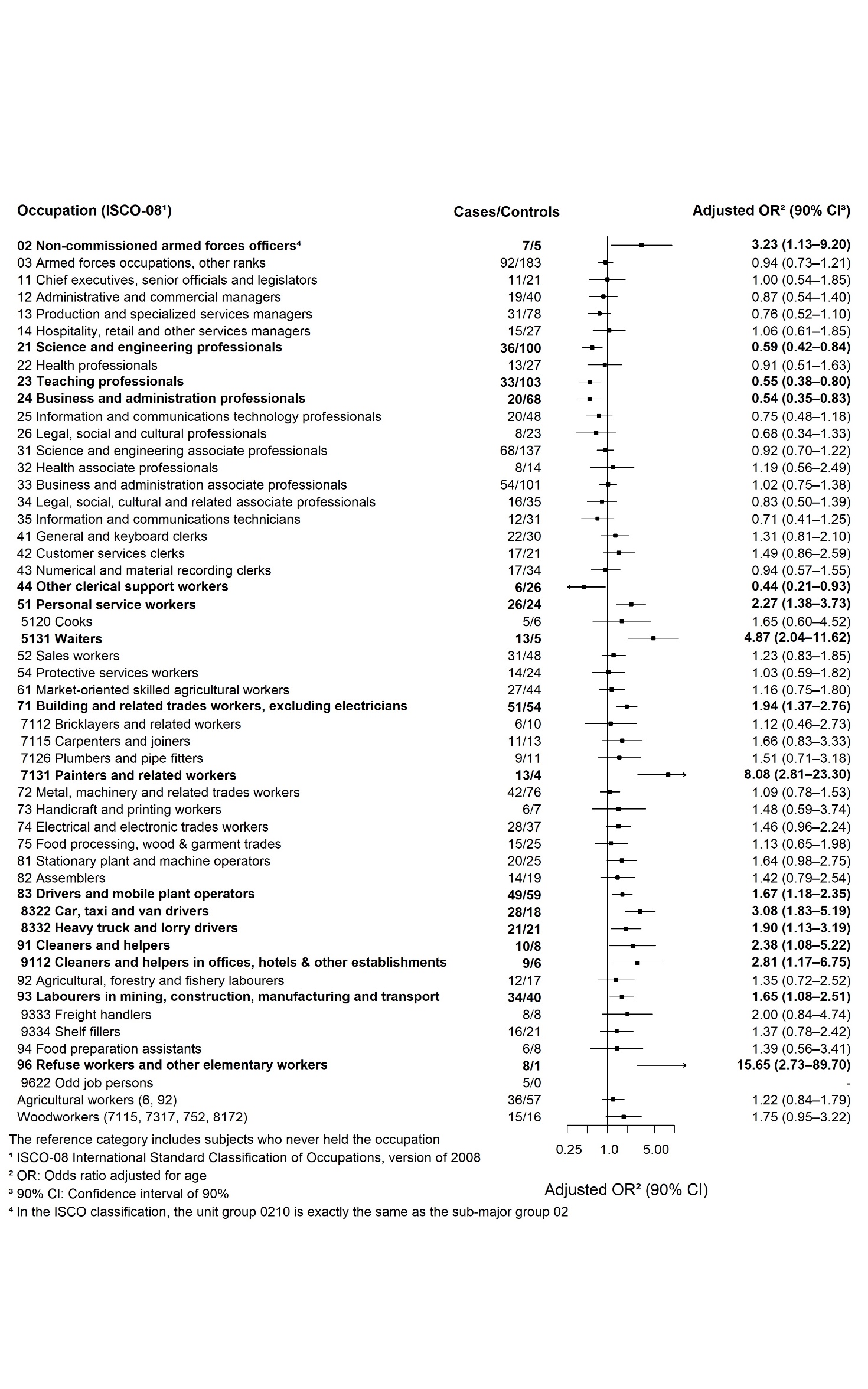

Figure S3: Odds ratios and 90% confidence intervals (adjusted for age) for the association between occupations (with at least 5 cases) and soft tissue sarcomas among men (336 cases / 626 controls) – ETIOSARC study, France, 2019-2023

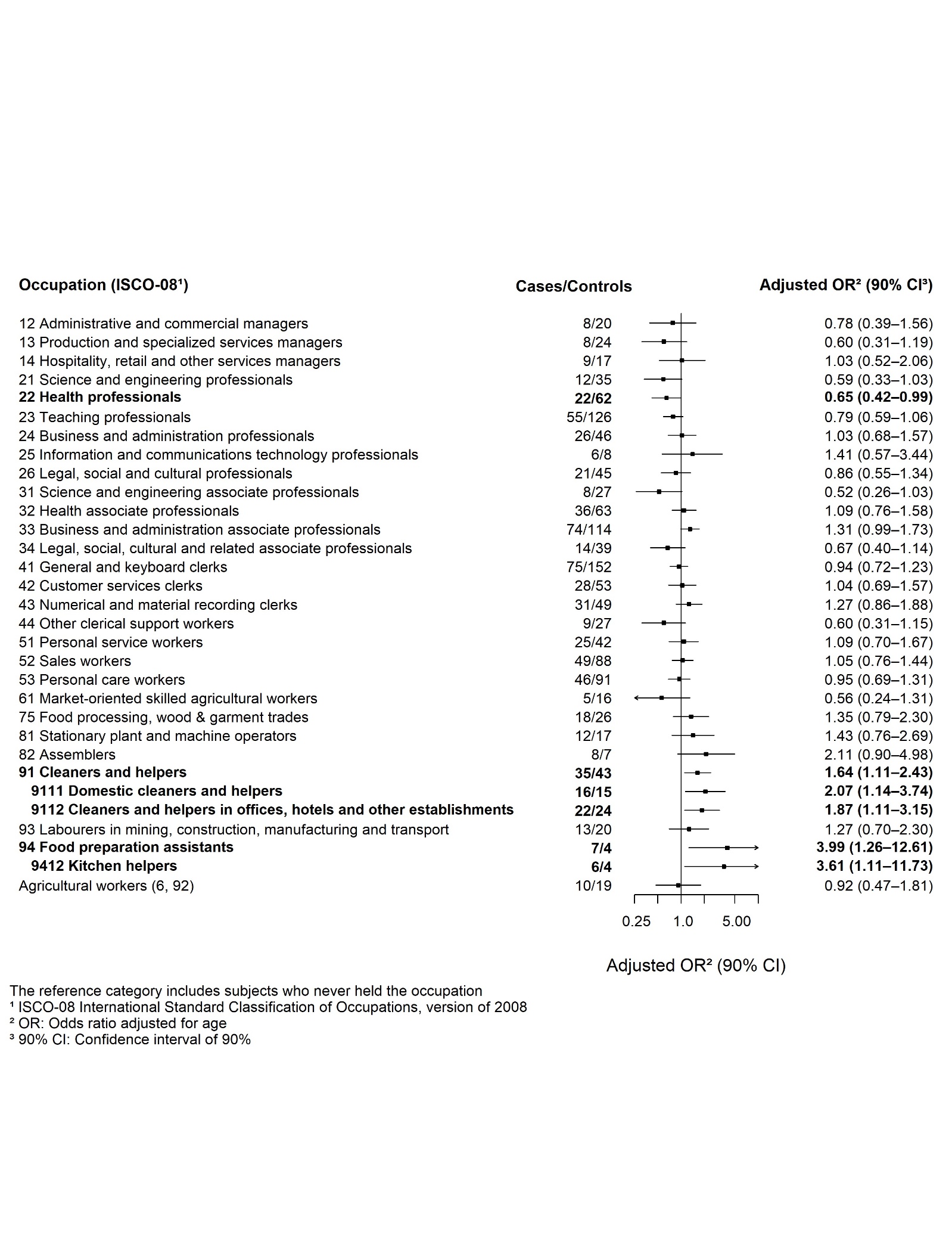

Figure S4: Odds ratios and 90% confidence intervals (adjusted for age) for the association between occupations (with at least 5 cases) and soft tissue sarcomas among women (335 cases and 631 controls) – ETIOSARC study, France, 2019-2023

Table S2: Odds ratios and 90% confidence intervals for the association between occupations (with at least 5 cases and OR > 1.5) and soft tissue sarcomas among men and women according to employment duration – ETIOSARC study, France, 2019-2023

|  | **< 5 years employed** | |  | **≥ 5 years employed** | |  |
| --- | --- | --- | --- | --- | --- | --- |
| **Occupations: ISCO-08^1^** | **Ca/Co^2^** | **OR^3^ (90% CI^4^)** |  | **Ca/Co^2^** | **OR^3^ (90% CI^4^)** | **p-value** |
| **Men** |  |  |  |  |  |  |
| 02 Non-commissioned armed forces officers | 3/3 | 2.79 (0.69-11.28) |  | 4/2 | 5.51 (1.26-24.05) | **0.09** |
| 51 Personal service workers | 15/9 | 3.38 (1.60-7.10) |  | 11/15 | 1.23 (0.62-2.41) | **0.02** |
| 5131 Waiters | 10/4 | 4.59 (1.70-12.43) |  | 3/1 | 4.34 (0.64-29.28) | **0.01** |
| 7131 Painters and related workers | 5/2 | 3.53 (0.88-14.14) |  | 8/2 | 8.96 (1.53-52.51) | **0.01** |
| 91 Cleaners and helpers | 7/7 | 1.70 (0.68-4.23) |  | 3/1 | 4.56 (0.67-31.12) | 0.24 |
| 96 Refuse workers and other elementary workers | 5/1 | 8.29 (1.36-50.46) |  | 3/0 | - | **0.01** |
| **Women** |  |  |  |  |  |  |
| 25 Information and communications technology professionals | 2/2 | 2.11 (0.40-11.11) |  | 4/6 | 1.47 (0.50-4.29) | 0.65 |
| 3334 Real estate agents and property managers | 4/0 | - |  | 1/3 | 0.79 (0.12-5.32) | **0.02** |
| 3343 Administrative and executive secretaries | 9/10 | 2.07 (0.94-4.57) |  | 18/15 | 2.57 (1.39-4.76) | **0.01** |
| 3344 Medical secretaries | 3/2 | 3.34 (0.73-15.27) |  | 8/10 | 1.52 (0.69-3.35) | 0.29 |
| 82 Assemblers | 6/6 | 1.48 (0.56-3.92) |  | 2/1 | 3.39 (0.45-25.51) | 0.47 |
| 94 Food preparation assistants | 4/0 | - |  | 3/4 | 1.30 (0.32-5.30) | **0.02** |
| 9412 Kitchen helpers | 3/0 | - |  | 3/4 | 1.31 (0.32-5.34) | **0.05** |

The reference category includes subjects who have never held the occupation

^1^ ISCO-08: International standard classification of occupations, version of 2008; ^2^ Ca/Co: Cases/Controls ever employed in the occupation; ^3^ OR: Odds ratio adjusted for age and education level; ^4^ 90% CI: confidence interval of 90%

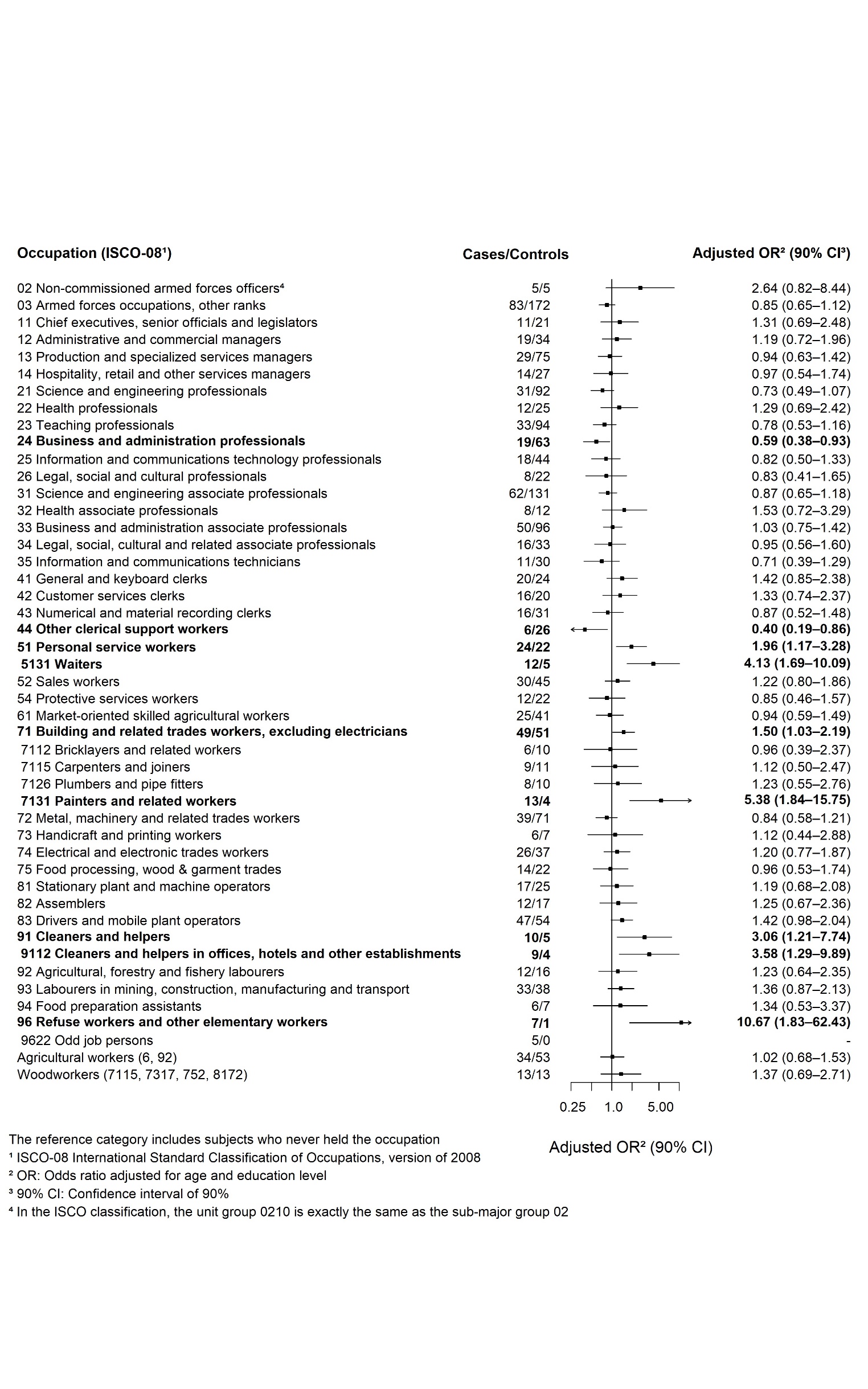

Figure S5: Odds ratios and 90% confidence intervals (adjusted for age and education level) for the association between occupations (with at least 5 cases) and soft tissue sarcomas among men (315 **histologically confirmed cases** and 586 controls) – ETIOSARC study, France, 2019-2023.

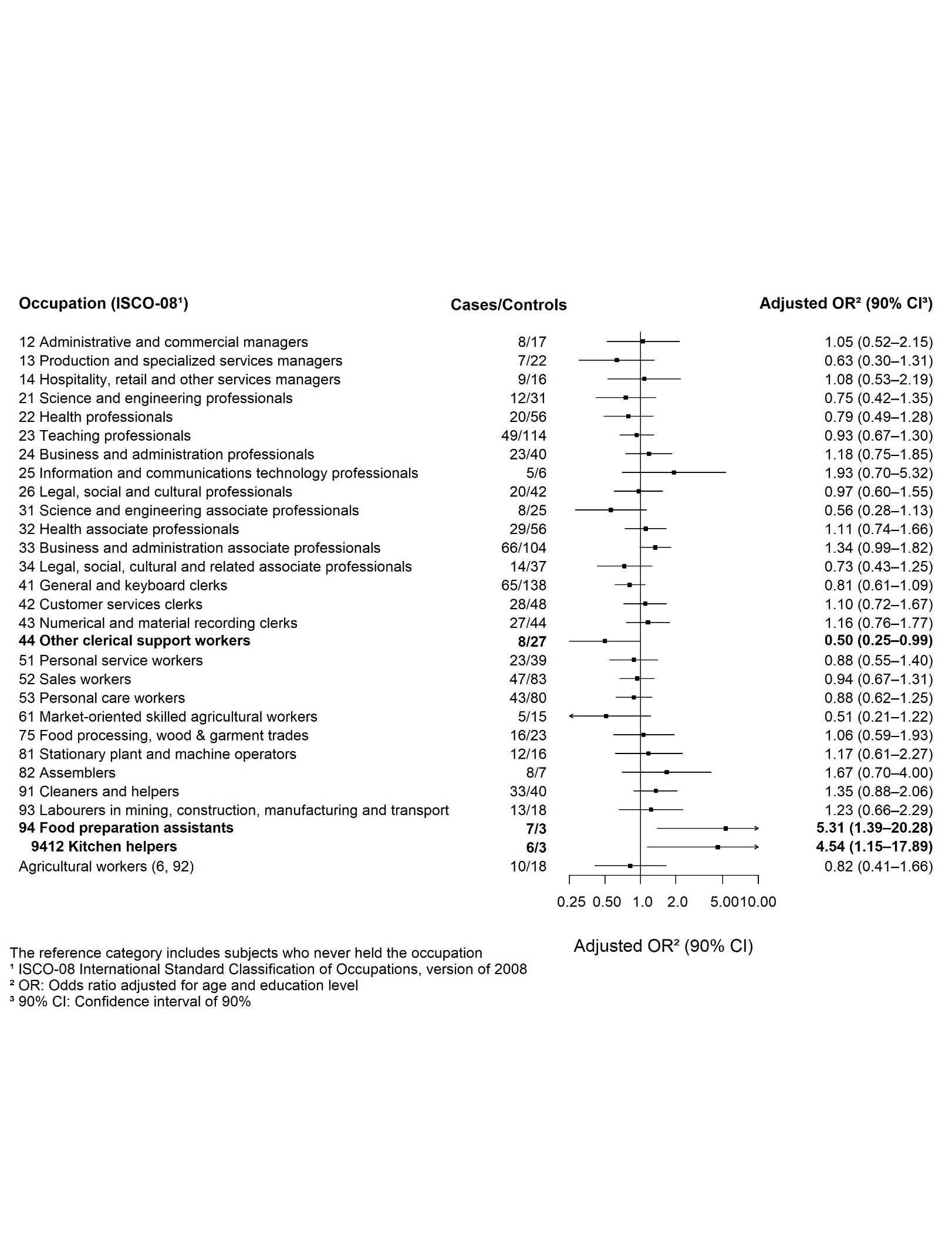

Figure S6: Odds ratios and 90% confidence intervals (adjusted for age and education level) for the association between occupations (with at least 5 cases) and soft tissue sarcomas among women (304 **histologically confirmed cases** and 570 controls) – ETIOSARC study, France, 2019-2023

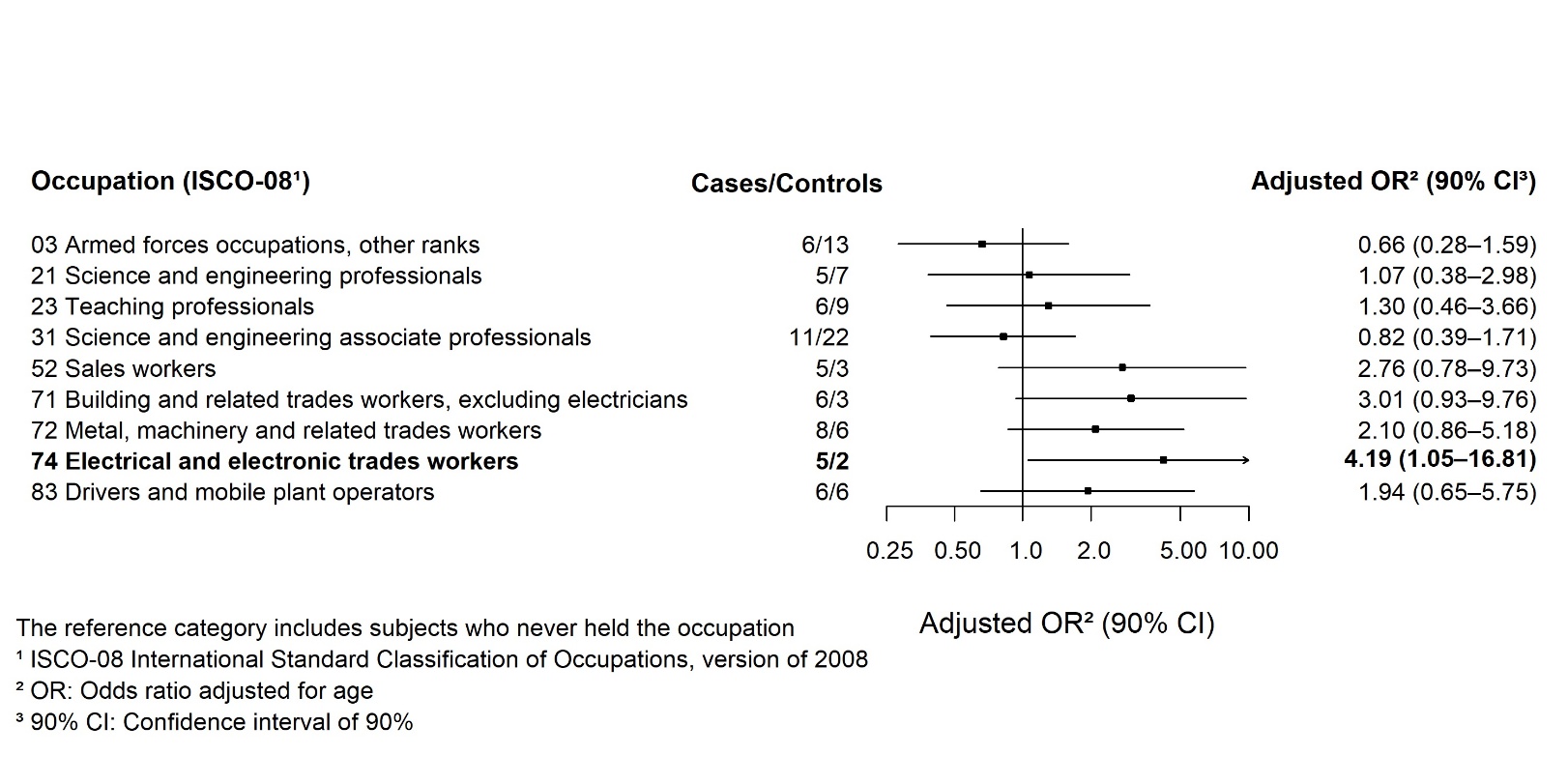

Figure S7: Odds ratios and 90% confidence intervals (adjusted for age) for the association between occupations (with at least 5 cases) and bone sarcomas among men (38 cases / 62 controls) – ETIOSARC study, France, 2019-2023

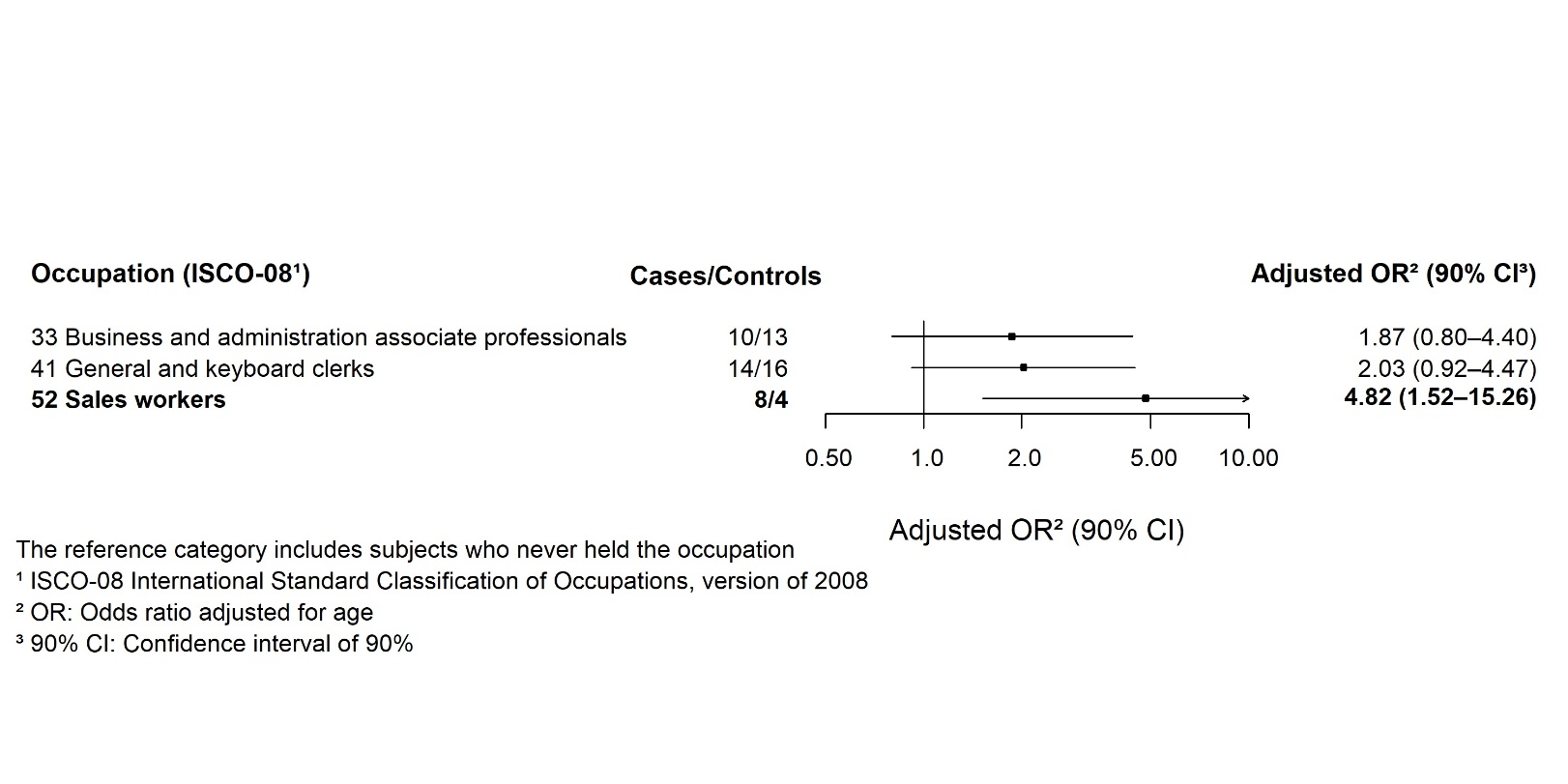

Figure S8: Odds ratios and 90% confidence intervals (adjusted for age) for the association between occupations (with at least 5 cases) and bone sarcomas among women (36 cases and 68 controls) – ETIOSARC study, France, 2019-202

Table S3: Odds ratios and 90% confidence intervals for the association between occupations (with at least 5 cases and OR > 1.5) and bone sarcomas among men and women according to employment duration – ETIOSARC study, France, 2019-2023

|  | **< 5 years employed** | |  | **≥ 5 years employed** | |  |
| --- | --- | --- | --- | --- | --- | --- |
| **Occupations: ISCO-08^1^** | **Ca/Co^2^** | **OR^3^ (90% CI^4^)** |  | **Ca/Co^2^** | **OR^3^ (90% CI^4^)** | **p-value** |
| **Men** |  |  |  |  |  |  |
| 21 Science and engineering professionals | 2/4 | 1.06 (0.20-5.61) |  | 3/3 | 2.18 (0.49-9.66) | 0.69 |
| 23 Teaching professionals | 2/6 | 1.13 (0.24-5.41) |  | 4/3 | 3.64 (0.89-14.89) | 0.30 |
| 52 Sales workers | 3/3 | 1.50 (0.35-6.44) |  | 2/0 | - | 0.14 |
| 71 Building and related trades workers, excluding electricians | 3/2 | 1.82 (0.37-8.96) |  | 3/1 | 3.36 (0.46-24.55) | 0.48 |
| 72 [Metal, machinery and related trades workers](javascript:__doPostBack('ctl00$ContentPlaceHolder1$TV_Code','sISCO2008\\425\\446')) | 4/2 | 2.62 (0.58-11.89) |  | 4/4 | 1.23 (0.36-4.27) | 0.54 |
| 74 Electrical and electronic trades workers | 2/2 | 1.59 (0.30-8.40) |  | 3/0 | - | **0.08** |
| 83 Drivers and mobile plant operators | 3/4 | 1.44 (0.32-6.45) |  | 3/2 | 1.66 (0.32-8.72) | 0.83 |
| **Women** |  |  |  |  |  |  |
| 33 Business and administration associate professionals | 5/3 | 7.48 (1.55-36.17) |  | 5/10 | 0.76 (0.24-2.48) | **0.07** |
| 41 General and keyboard clerks | 5/4 | 2.02 (0.61-6.66) |  | 9/12 | 1.48 (0.55-3.98) | 0.56 |
| 52 Sales workers | 6/1 | 19.99 (2.84-140.52) |  | 2/3 | 0.99 (0.16-6.06) | **0.01** |

The reference category includes subjects who have never held the occupation

^1^ ISCO-08: International standard classification of occupations, version of 2008; ^2^ Ca/Co: Cases/Controls ever employed in the occupation; ^3^ OR: Odds ratio adjusted for age and education level; ^4^ 90% CI: confidence interval of 90%

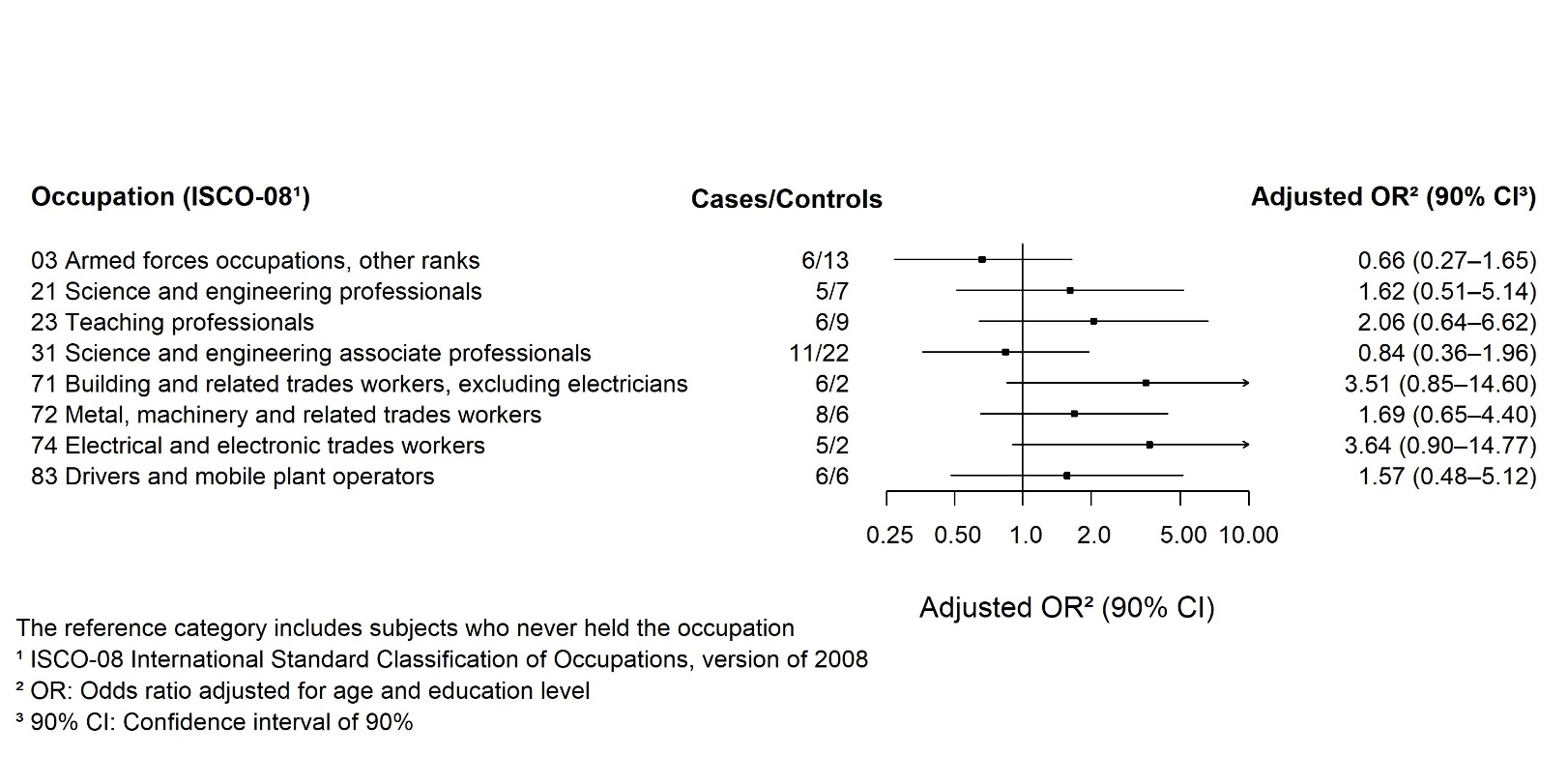

Figure S9: Odds ratios and 90% confidence intervals for the association between occupations (with at least 5 cases) and bone sarcomas among men (37 **histologically confirmed cases** and 60 controls) – ETIOSARC study, France, 2019-2023

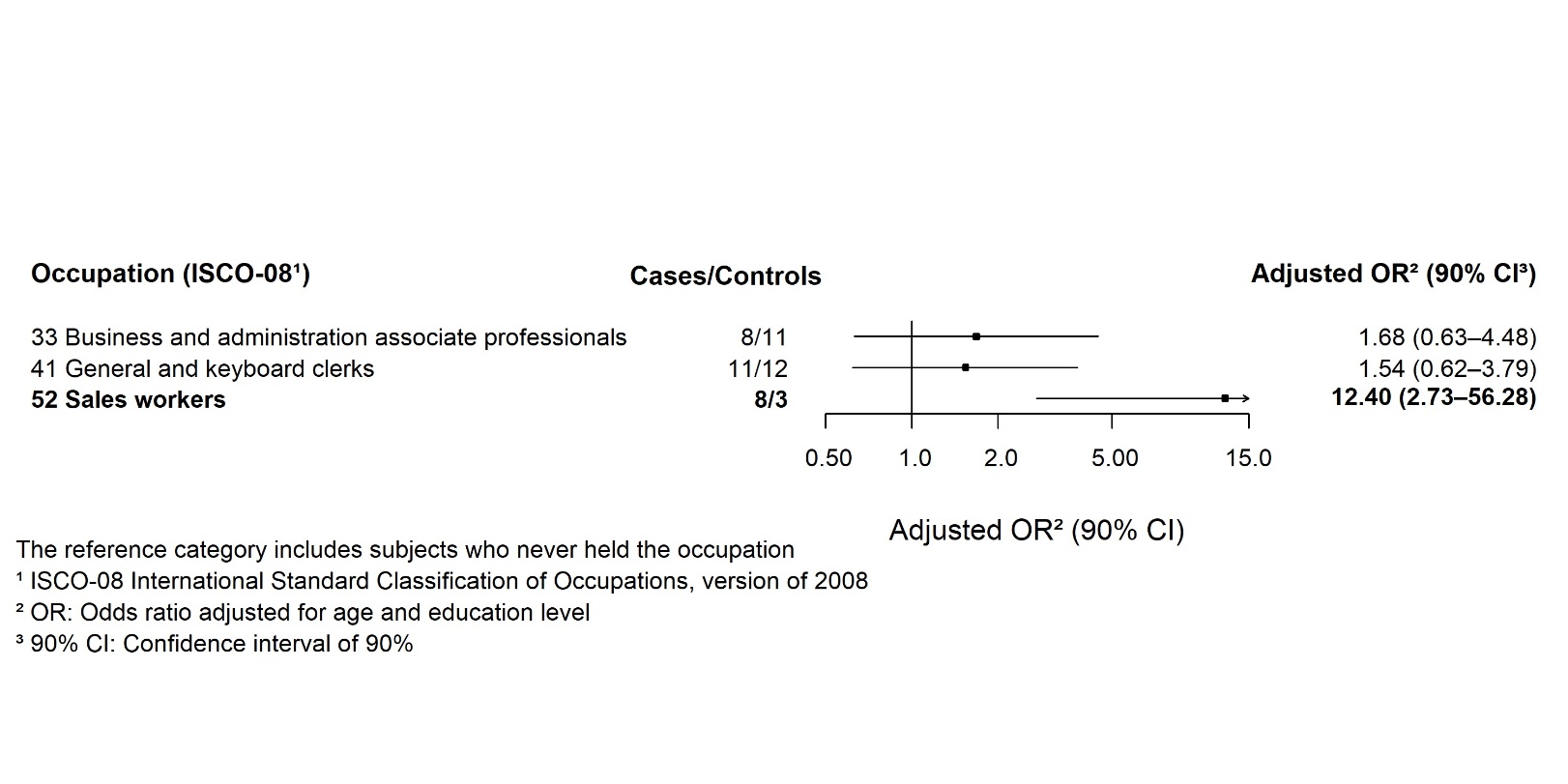

Figure S10: Odds ratios and 90% confidence intervals for the association between occupations (with at least 5 cases) and bone sarcomas among women (30 **histologically confirmed cases** and 56 controls) – ETIOSARC study, France, 2019-2023
